# Cross Validation Assessment of Clinical Performance Characteristics of a Test Table for the Risk of Maternal Autoantibody-Related Autism in High-Risk Populations

**DOI:** 10.64898/2026.10.23.26366366

**Authors:** E. Robert Wassman, Megan Martin, Joseph Schauer, Judy Van de Water, Michael Paul

## Abstract

**Background/Objectives:** Specific combinations of maternal autoantibodies targeting fetal brain proteins are associated with Maternal Autoantibody-Related Autism (MARA), a biologically defined subset of autism. We previously translated a multi-ELISA assay to detect MARA-associated autoantibodies and identify mothers with transcendent levels of specific autoantibody combinations that exceed that of controls and are highly predictive of autism in their offspring. Our objective here was to derive a rule-based classifier, a “test table” of autoantibody pairwise combinations, each with pair-specific thresholds, and to validate its clinical performance characteristics by cross-validation.

**Methods:** Plasma samples from mothers of 482 autism and 425 typically developing children were analyzed in a clinical lab for previously identified predictive combinations of autoantibodies. Stratified 20-fold cross-validation in all 907 participants assessed test table stability and predictive performance. The most stable test table was retrained on the full cohort to establish optimal thresholds and determine performance. Permutation testing assessed performance relative to chance.

**Results:** Cross-validated performance showed a mean specificity across folds of 99.8%, an imputed mean PPV of 97.5%, a detection rate of 4.4%, and a positive likelihood ratio (LR+) of 18.5. Applied in-sample to the full cohort, the final test table detected 6.2% of cases with no false positives, giving an upper-range LR+ estimate of 26.5.

**Conclusion:** The high specificity, PPV, and positive likelihood ratio of this assay support its potential clinical implementation for identifying a subset of mothers at very high risk of having a child with autism, and as a companion diagnostic for interventions targeting the MARA subtype of autism.

## Introduction

Autism is an important public health concern and a major socioeconomic challenge for affected individuals, their families, and the healthcare system, with recent data showing 1 in 31 children in the United States are affected [1,2]. A neurodevelopmental disorder characterized by varying difficulties with social interaction and communication along with restricted or repetitive patterns of thought and behavior, autism has its onset in the early years of life, supporting the importance of prenatal influences [1]. While genetics and environmental factors are drivers of the etiology of autism, the maternal immune system during pregnancy has been shown to have a significant role in many affected individuals [3–5].

Autism is diagnosed by comprehensive clinical and psychological evaluation, which is highly accurate but resource-intensive. Long wait times for these definitive diagnostic evaluations, typically until between three and five years of age, results in delayed access to early intervention and support [6]. Biomarkers linked to the underlying biology of autism could provide a critical clinical tool to facilitate earlier diagnosis and, if mechanistically involved in its etiology, may also identify targets for prevention or mitigation.

The Van de Water laboratory at the MIND Institute at the University of California, Davis first described maternal autoantibodies targeting fetal brain proteins critical to neurodevelopment in 2008 [7]. It is well known that maternal IgG antibodies routinely traverse the placenta via the neonatal fragment crystallizable receptor (FcRn); likewise, pathological maternal autoantibodies can cross as well, causing deleterious effects in the fetus/newborn child by damage to their fetal antigenic targets [8–11]. Specific combinations of MARA autoantibodies which target developing brain proteins can access antigens within the developing brain at critical periods of fetal brain development when the blood brain barrier is not yet fully functional [10,11], and these have been shown to correlate with the development of a subset of autism [4,7,12,13]. Animal models in mice, rats and sub-human primates support the pathogenic role of these MARA autoantibodies [1,14–19]. Both passive antibody transfer into dams or the active induction of maternal autoantibody production in animal models resulted in neuroanatomical alterations and autistic-like behavioral phenotypes in the offspring following gestational exposure to these autoantibodies [1,14–19]. Thus, MARA autoantibodies represent mechanistic biomarkers which epidemiological studies suggest are potentially useful indicators of autism risk.

MARA autoantibody combinations targeting specific pairs of eight proteins that are highly expressed during brain development have been observed in between 10% and 24% of mothers of children with autism across several epidemiologic studies [4,7,12,13,20–23]. Children of mothers with these autoantibodies tend to show greater autism symptom severity. The prospective Early Markers for Autism (EMA) study demonstrated that these potentially pathogenic autoantibodies were present during pregnancy, before the birth of the first affected child [13]. Analyses of these autoantibodies in mothers of children enrolled in the CHARGE (The Childhood Autism Risks from the Genetics and Environment) study, a large case–control study investigating potential causes of autism (genetic or environmental), provided further evidence of the potential significance of these autoantibodies in the etiology and prediction of autism [24]

MARA autism-specific patterns in the CHARGE cohort demonstrated greater than 99% classification accuracy and identified the likelihood of subsequent diagnosis of autism in the children enrolled in the study [7,13,20,25]. Recent comprehensive reviews of the maternal autoantibody correlation to neurodevelopmental disorders summarized extensive evidence that maternal autoantibodies reactive to specific fetal brain proteins may be a highly specific biomarker of autism risk, and they are potentially modifiable environmental contributors to development of autism [26,27].

Rigorous translation of these mechanistic biomarkers into clinical application provides an important avenue toward earlier identification of autism and, potentially, the development of targeted therapeutic or preventive interventions for a subset of autistic individuals. Recently we reported the analytical validation to clinical standards of enzyme-linked immunosorbent assays (ELISAs) for detection of the eight previously reported MARA autoantibodies [(Collapsin response mediator proteins 1 and 2 (CRMP-1 and CRMP-2), Guanine deaminase (GAH), Stress-induced phosphoprotein 1 (STI1), Neuron-specific enolase (NSE), L-lactate dehydrogenase A and B chains (LDH-A and LDH-B), and Y-box-binding protein 1 (YB-1)] [28]. Following validation of this set of MARA autoantibodies in two independent Clinical Laboratories Improvement Act (CLIA) certified laboratories, we conducted a staged clinical evaluation of the correlation of the combined multi-ELISA assay with autism. In the first part of these studies (reported elsewhere [30]), de novo discovery conducted on the CHARGE cohort [24] identified a “transcendent” autoantibody profile in a subset of mothers of autistic children (70/420; 16.7%) whose levels of both autoantibodies in at least one combination exceeded those of every control mother in the cohort. Because no control mother can meet the transcendent criteria, specificity and positive predictive value (PPV) for a transcendent result are 100% by construction. Transcendence was then replicated in a small external cohort [29] from the Markers of Autism Risk in Babies—Learning Early Signs (MARBLES) study (n = 132), an independent prospective longitudinal study, which followed women with a previous child diagnosed with autism who were pregnant or planning a subsequent pregnancy [30]. This seminal observation using an analytically validated assay confirmed the results from earlier published academic studies that used a non-clinical prototype eight-autoantibody assay to detect the MARA subset of autism [13,20]. This supports the potential for highly specific discrimination between children with autism and all typically developing children, and is consistent with the hypothesis that the assay detects a MARA subset of autism.

The objective of this study was to develop and estimate the clinical performance of a rule-based classifier, or “test table”, suitable for clinical use in high-risk intended use populations. The intended use is to identify individuals for whom a positive result would indicate a very high post-test probability of autism, rather than broad population screening utility. In this framework, the classifier developed here is not a conventional continuous risk score; instead, it is a rule-based table of autoantibody pairs and pair-specific thresholds, with positivity defined by satisfying at least one pairwise rule. In this study, we report performance metrics derived from stratified 20-fold cross-validation rather than apparent in-sample performance [31,32]. In cross-validation, the cohort is repeatedly split so that each participant is tested by a version of the test table developed without that participant. This reduces the overly optimistic results caused by overfitting, which occurs when a classifier is tested on the same data used to build it. The final test table was then built using the full cohort. Performance estimates are reported from cross-validation, and the apparent (in sample) performance of the final test table is also reported.

## Materials and Methods

### Previous Analyses and Present Study Design

Banked maternal plasma samples were assayed in a CLIA/CAP-certified laboratory for eight autoantibodies to: Collapsin response mediator proteins 1 and 2 (CRMP-1 and CRMP-2), Guanine deaminase (GAH), Stress-induced phosphoprotein 1 (STI1), Neuron-specific enolase (NSE), L-lactate dehydrogenase A and B chains (LDH-A and LDH-B), and Y-box-binding protein 1 (YB-1), all previously extensively described as MARA autoantibody antigens [4,7,12,13,20,21,23,28,29].

Our prior analyses of the CHARGE cohort (420 mothers of autistic children, 335 mothers of typically developing (TD) children) [29] produced three findings that shaped the design of the present study. First, 70 mothers of autistic children (16.7%) were transcendent: their levels of both autoantibodies in at least one of the 28 possible combinations exceeded those of every control mother. The number of transcendent cases was significantly greater than expected by chance by permutation testing. This separation from controls informed how test table thresholds were relative to the upper limit of control values. Second, transcendence was concentrated in a few combinations. Five (CRMP-1 + CRMP-2, LDH-A + STI1, LDH-B + YB-1, STI1 + YB-1, and CRMP-2 + YB-1) accounted for 60 of the 70 transcendent cases, suggesting that a small panel of combinations could capture most of the high-risk cases. Third, transcendence was replicated in the independent MARBLES cohort (42 mothers of autistic children, 90 mothers of TD children) showing a similar bimodal pattern of risk scores. Fourteen mothers (33.3%) were transcendent, significantly more than expected by chance. Nine of these were identified by the same five combinations, which supported pooling the two cohorts for test table development and cross-validation.

Building on this discovery and external validation work, the present study developed the test table using the union of the CHARGE and MARBLES cohorts for a total of 907 participants of which 482 had children with autism, and 425 TD. The study proceeded in four steps. First, starting with 15 autoantibody combinations, stratified 20-fold cross-validation was used to estimate the performance and stability of the test table development procedure. In each fold, the full procedure (combination selection, threshold derivation, and threshold inflation) was performed on the training partition only, and the resulting test table was applied to the held-out participants. Second, the same procedure was applied once to all 907 participants to produce the final test table, and its apparent (in-sample) performance was determined. Third, permutation testing was used to determine whether the number of selected combinations and test-positive autism cases exceeded what would be expected by chance: the development procedure was repeated 10,000 times with autism and TD labels randomly reassigned. Fourth, a risk analysis examined the distribution of autism and TD participants across levels of predicted risk, derived by progressively lowering the thresholds of the final test table. Performance metrics of the final test table was derived exclusively from cross-validation.

### Specimen Sources

907 Bio-banked acid–citrate–dextrose (ACD) plasma samples from the CHARGE and MARBLES studies were obtained from mothers of 482 autistic and 425 typically developing (TD) children as previously described [29]. Participants were originally selected based on the inclusion/exclusion criteria of the CHARGE and MARBLES studies, and samples were collected, processed and stored as described in the respective protocols [24,30].

The subject inclusion criteria for CHARGE was mothers of children diagnosed with autism by gold standard clinical diagnostic protocols at the MIND Institute/UC Davis, while MARBLES inclusion criteria was women with one or more biological children previously diagnosed with autism by the same clinical criteria, who were pregnant or planning a future pregnancy. Controls from CHARGE were randomly selected from birth records, frequency matched on age, sex, and broad geographic region, and confirmed as neurotypical (typically developing; TD) based on the Social Communication Questionnaire (SCQ) [33]. All available 24-month post-partum maternal samples following a subsequent child’s birth were analyzed from MARBLES [29]. There were three subjects from CHARGE that also appeared in this subcohort of MARBLES.

Only biobanked patient samples were used in this study. Both the previously conducted CHARGE and MARBLES studies complied with all applicable regulatory requirements regarding human participants, including informed consent obtained before collection of any data, and approval by the institutional review boards for the State of California and the University of California [24,30].

### Blinding

De-identified study samples were assayed blindly at the CLIA laboratory. Permutation analysis and cross-validation data were unblinded as the randomization steps in these processes obviate the need for blinding. MARAbio Clinical, Development, and Research personnel were blinded to the clinical validation study results until the analysis was completed.

### MARA Autoantibody ELISA Procedure Methodology

All maternal blood samples had been collected in 8 ml ACD tubes (BD Diagnostics) and plasma was separated, labeled, aliquoted, and stored at −80 °C within 24 hours of drawing. Banked samples from CHARGE and MARBLES that had been frozen and thawed more than twice were excluded.

The ELISA procedure methodology was performed as described previously in detail [27]. Briefly, all assays were indirect ELISAs with recombinant protein antigens (LDH-A, LDH-B, GAH, STI1, CRMP-1, CRMP-2, NSE, or YB-1) (sourced from Expression Systems, LLC, Davis, CA, USA) bound to microarray plates, incubated with the test maternal plasma samples, Goat anti-human IgG horseradish peroxidase (HRP)-conjugated secondary antibody, and finally a chromogenic substrate prior to reading optical density (OD) on a spectrophotometer at 450 nm with a reference at 595 nm. A single point calibration method was used along with a reagent blank control. In positive samples, blue color developed in the wells at an intensity proportional to the concentration of protein-specific antibodies.

### Test Table Development and Cross-Validation Determination of Stability

20-fold cross-validation [31,34,35] was performed on the combined CHARGE and MARBLES cohorts (n=907) to assess the stability of the MARA test table and estimate its predictive performance. Cross-validation was used because performance metrics estimated from the same cohort used to develop an assay will typically overestimate the performance of an assay. Although we previously conducted a small external validation study [29] to confirm the key findings and support the potential performance of the assay, the cross-validation performed in this study provides an estimate of the performance that might be expected in a larger independent validation cohort, i.e., as yet unseen participants.

The derived classifier was structured as a test table defined by a set of autoantibody pairs (combinations), each defined by two autoantibodies (P1, P2) and corresponding concentration thresholds (T1, T2, in ELISA units (EU)). A sample was classified as test-positive if for any combination in the test table the sample’s assay values for P1 and P2 both met or exceeded T1 and T2 respectively; otherwise, the sample was test-negative. Importantly, the characteristics of a good test table included stability of the autoantibodies selected for inclusion in the test table, high specificity (≥99%) and high positive predictive value (PPV) (≥95%). We note that the PPV was assessed at the study prevalence which was 52% (CHARGE + MARBLES). The term ‘detection rate’ refers to the percentage of true positive autism cases detected, which approximates sensitivity.

A test table was considered stable when after cross-validation, the same autoantibody pairs were selected as being most predictive. Thus, following completion of cross-validation, the most stable test table was selected and test table thresholds were recalibrated on the full 907-participant dataset to produce the final test table. Folds were assigned at random with stratification on case status, preserving the cohort case-control ratio across folds. All threshold calibration steps were confined to the training partition of each fold [31]. Twenty folds were prespecified so that each training partition contained 95% of participants, minimizing the pessimistic bias of k-fold estimates relative to the final model trained on the full cohort [31,36].

Within each training fold, candidate autoantibody pairs were evaluated against prespecified inclusion criteria (specificity of at least 99.5%, PPV at least 95%) on the training partition. NSE and GAH were excluded from final test table development because no combinations involving these autoantibodies were observed in cross-validation **(Figure 1)** and none in transcendents in our prior discovery analysis [29]; candidate combinations were therefore drawn from the 15 pairwise combinations of the remaining six autoantibodies. To reduce the risk that thresholds derived in training would generate false positives on the held-out data, threshold values were inflated prior to lock by 3% for T1 and 5% for T2, multiplied directly onto the log scale threshold values.

**Figure 1.**
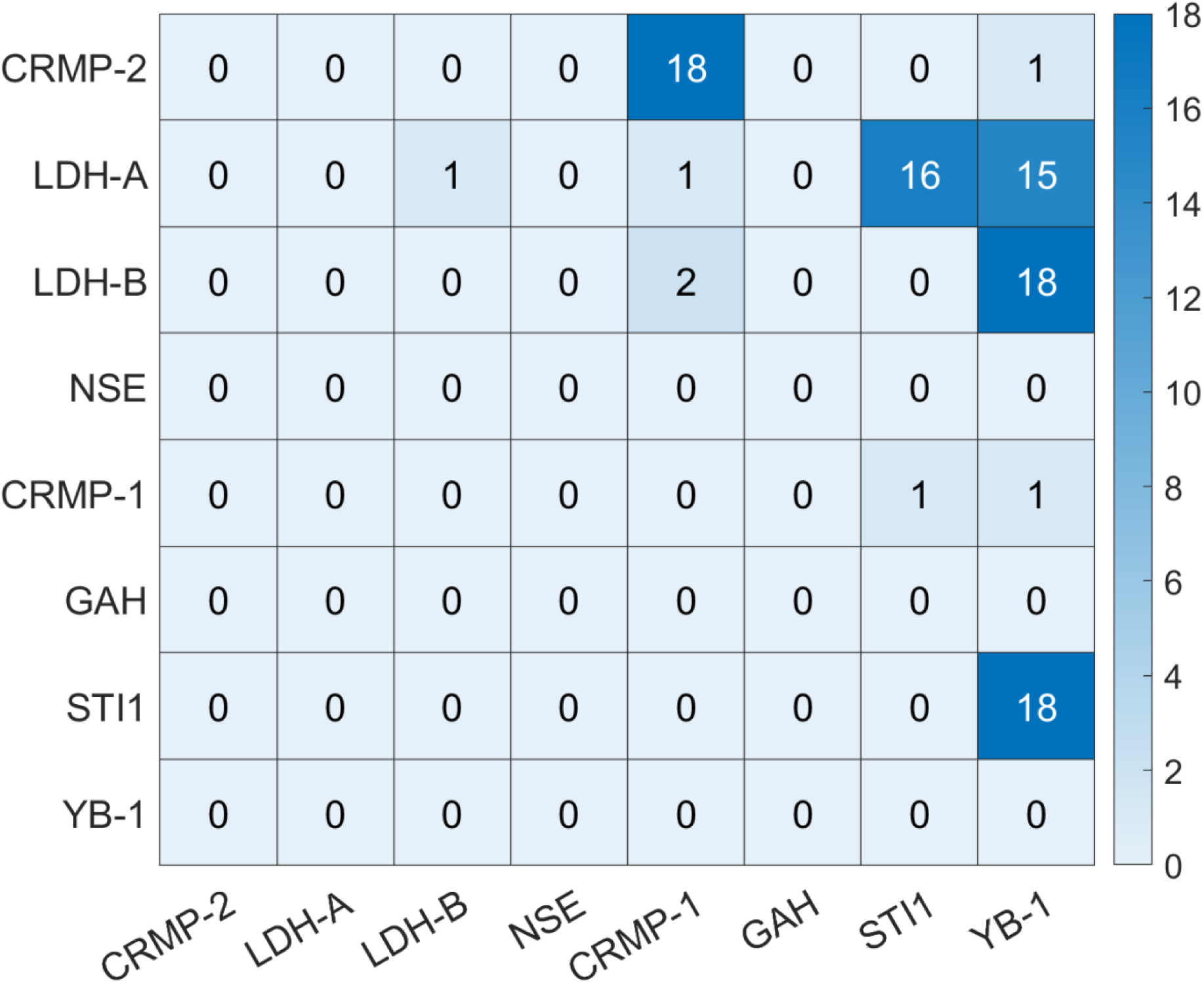
Reproducibility of autoantibody pairs across 20 cross-validation folds. Each cell represents a pairwise combination of the autoantibodies listed on the row and column axes. The number in each cell is the number of folds (0 to 20) in which that combination was selected for the test table developed on the training partition. Darker shading indicates more frequent selection.

Folds 1 through 19 each held out 25 cases and 22 controls; fold 20 held out the remaining 7 cases and 7 controls. The full training procedure was repeated independently within each fold; the resulting test table was applied to the held-out partition; and fold-level true positives, false positives, sensitivity, specificity, and PPV were recorded. For each fold, the entire threshold recalibration pipeline was re-run de novo, the resulting test table was then applied to the held-out fold, and fold-level true positives, false positives, sensitivity, specificity, and PPV were recorded.

Final performance estimates were determined from cross validation alone. Because thresholds were inflated in training, cross-validation estimates are conservative with respect to specificity (biased upwards) and sensitivity (biased downwards) relative to a non-inflated estimator. This conservative bias was a deliberate choice consistent with the test’s intended use as a high-specificity rule-in test.

### Statistical analysis

Mean specificity across folds was reported with a 95% confidence interval calculated using the Nadeau and Bengio correction, which accounts for the non-independence of training partitions across folds [37]. Because some held-out folds contained no positive test calls, fold-level PPV was undefined when true positives (TP) + false positives (FP) = 0. In this event two PPV summaries were therefore reported. The raw summary was computed only over folds with at least one positive call. The imputed summary additionally assigned 100% PPV to folds in which the test made no positive calls. In these folds the empirical PPV was undefined, but no false positives occurred, an outcome consistent with the intended interpretation of a high-specificity screen in very small test folds.

The fold-level sensitivity is expected to underestimate the true sensitivity due to the cohort’s low expected positives per fold. Several folds were expected to produce zero positives by chance alone, even when the underlying classifier has a nonzero detection rate. Consequently, at the end of the procedure, in-sample sensitivity was calculated directly.

Following completion of initial cross-validation, test table thresholds were retrained on the full 907-participant dataset to generate the final test table and performance metrics derived [32]. The same procedure used within the cross-validation was used to derive the retrained test table. Consistent with good prediction-model reporting practice, the retrained test table was reported descriptively, whereas inference about expected future performance in an as yet unseen population is based on cross-validation results only.

The Fisher exact test was used to test for significance of association for both cohorts.

In addition to reporting stability and performance from cross validation, binomial testing was used to test if there were more test-positive autism cases than expected by chance alone. Although the binomial test is appropriate to determine if there is a significant cluster of high-risk autism subjects selected by the test table, permutation testing [38,39] was also performed to determine both the test table size (number of combinations) and number of high-risk autism cases obtained on random data. The permutation testing procedure estimated the frequency that a test table with the same or better performance characteristics (specificity, PPV, sensitivity) as the final test table would be observed across 10,000 iterations where subject labeling as autism or TD was randomized [40]. The probability of this event was then calculated as (k+1)/10001 where k is the observed number of events. The permutation test procedure utilized the same threshold retraining pipeline embedded within the cross validation described above. This was conducted on all 907 subjects in the combined cohort.

### Risk Analysis

Risk analysis was performed to assess the distribution of autism and TD participants with respect to predicted risk. Predicted risk was derived from the test table generated by the cross-validation procedure described above, by progressively scaling all thresholds in the test table downward (by factors of 1.00, 0.99, 0.98, …, 0.01) and recording the resulting true positive and false positive calls at each scaling level. At each percentage value the PPV value of the derivative test table was calculated as well as the relative proportion of test positive participants that have autism or are TD.

## Results

Results presented are of the assessment of stability and performance by cross validation on the combined CHARGE and MARBLES cohorts.

### Cross-validation stability

The 20-fold cross validation procedure identified five autoantibody combinations as stable selections across folds: CRMP-2 + CRMP-1, LDH-A + STI1, LDH-A + YB-1, LDH-B + YB-1, and STI1 + YB-1. All five combinations were selected in a majority of folds (15 to 18 out of 20 folds), whereas no other combination was selected in more than two folds **(Table 1**, **Figure 1).**

**Table 1.** Reproducible combinations selected across cross-validation folds.

| **Combinations** | **Fold Occurrence (out of 20)** |
| --- | --- |
| CRMP-2 + CRMP-1 | 18 |
| LDH-B + YB-1 | 18 |
| STI1 + YB-1 | 18 |
| LDH-A + STI1 | 16 |
| LDH-A + YB-1 | 15 |

### Final test table and performance metrics

Following cross-validation, the final test table was derived by model retraining, by applying the same test table selection procedure to all 907 participants in the combined cohort from CHARGE and MARBLES, using the same inclusion criteria and threshold inflation [32]. As the control set is small, we used artificially inflated decision thresholds to ensure that false positives would be extremely unlikely in an unknown clinical population.

The final test table **(Table 2)** contains the same five reproducible autoantibody combinations identified across the cross-validation folds **(Table 1)**, with thresholds derived from the full cohort. Other autoantibody combinations are not reported in the final test table as the frequency of true positives was negligible.

**Table 2.** Final retrained test table.

| **P1** | **P2** | **T1 (EU)** | **T2 (EU)** |
| --- | --- | --- | --- |
| CRMP-2 | CRMP-1 | 180.8 | 467.5 |
| LDH-A | STI1 | 266.4 | 148.3 |
| LDH-A | YB-1 | 93.1 | 927.0 |
| LDH-B | YB-1 | 145.9 | 215.7 |
| STI1 | YB-1 | 110.0 | 493.8 |

P1 and P2 indicate the two autoantibodies in each combination; T1 and T2 are their respective concentration thresholds in ELISA units (EU). A sample is classified as test positive if, for any row, the participant's assay concentrations of both P1 and P2 meet or exceed T1 and T2 respectively.

### Performance metrics from 20-fold cross-validation

Across the 20 held-out folds, raw PPV was 100% in 10 folds, 50% in 1fold (which contained 1 true positive and 1 false positive), and undefined in the remaining 9 folds because no positives were detected. However, in all 9 undefined folds, no false positives were generated either, so these folds were assigned a PPV of 100% for the imputed mean PPV calculation. Fold-level specificity was 100% in 19 of 20 folds and 95.5% in the single fold containing the false positive.

The performance metrics were determined after completion of the 20-fold cross-validation. The mean specificity across folds was 99.8% with a 95% confidence interval of 99.2% to 100.0%. The mean PPV among folds with reportable PPV was 95.5% (95% CI 84.7% to 100.0%), and the imputed mean PPV was 97.5% (95% CI 91.3% to 100.0%). When the final test table was applied to the full cohort, 30 true positives and no false positives were observed. The specificity, PPV, and NPV were 100%, 100% and 47.9% respectively by this in-sample assessment. **(Table 3)**

**Table 3.**
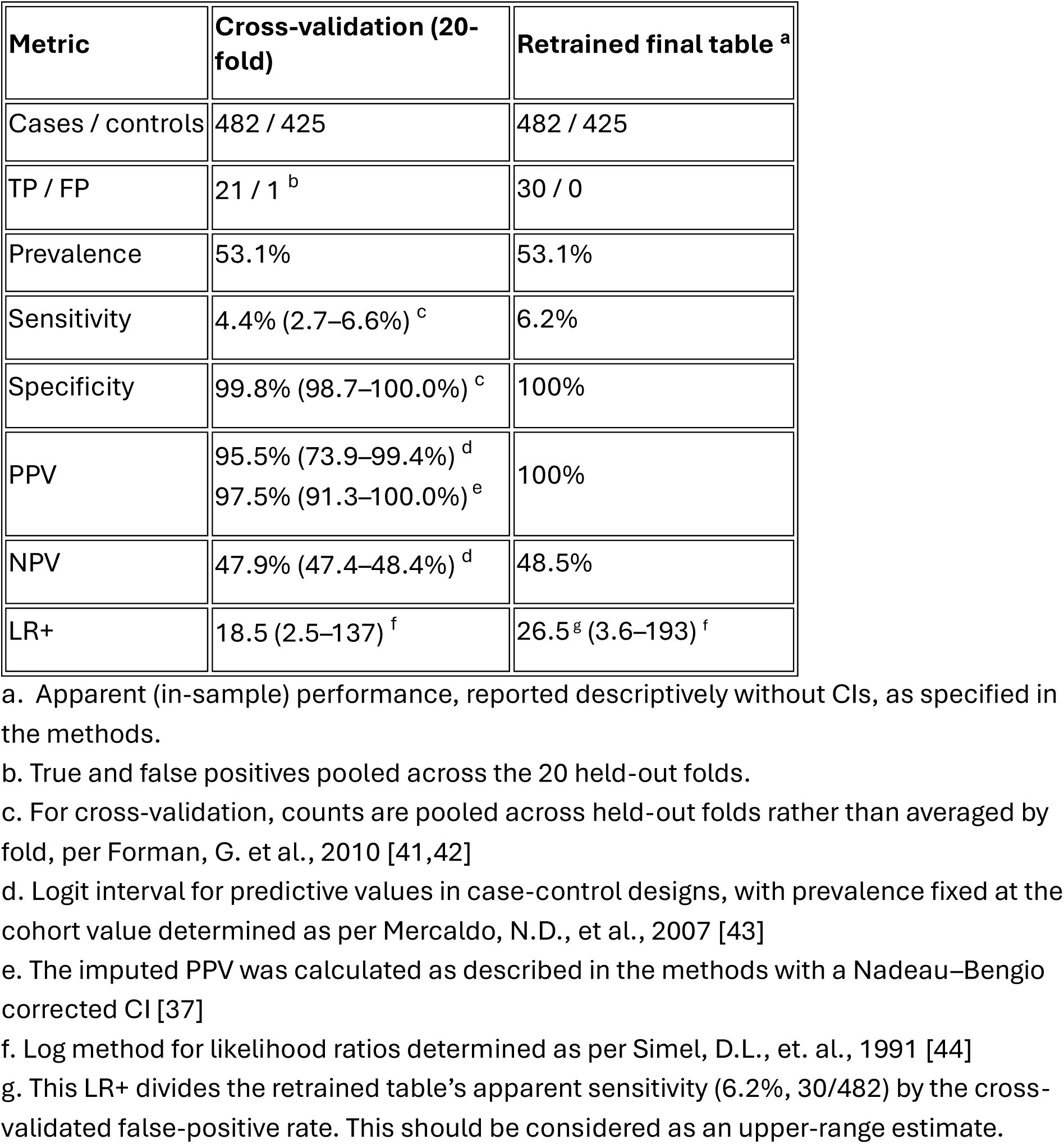
Estimation of performance by cross-validation and final test table in-sample.

| **Metric** | **Cross-validation (20-fold)** | **Retrained final table <sup>a</sup>** |
| --- | --- | --- |
| Cases / controls | 482 / 425 | 482 / 425 |
| TP / FP | 21 / 1 <sup>b</sup> | 30 / 0 |
| Prevalence | 53.1% | 53.1% |
| Sensitivity | 4.4% (2.7–6.6%) <sup>c</sup> | 6.2% |
| Specificity | 99.8% (98.7–100.0%) <sup>c</sup> | 100% |
| PPV | 95.5% (73.9–99.4%) <sup>d</sup><br>97.5% (91.3–100.0%) <sup>e</sup> | 100% |
| NPV | 47.9% (47.4–48.4%) <sup>d</sup> | 48.5% |
| LR+ | 18.5 (2.5–137) <sup>f</sup> | 26.5 <sup>g</sup> (3.6–193) <sup>f</sup> |

a. Apparent (in-sample) performance, reported descriptively without CIs, as specified in the methods.
b. True and false positives pooled across the 20 held-out folds.
c. For cross-validation, counts are pooled across held-out folds rather than averaged by fold, per Forman, G. et al., 2010 [41,42]
d. Logit interval for predictive values in case-control designs, with prevalence fixed at the cohort value determined as per Mercaldo, N.D., et al., 2007 [43]
e. The imputed PPV was calculated as described in the methods with a Nadeau–Bengio corrected CI [37]
f. Log method for likelihood ratios determined as per Simel, D.L., et. al., 1991 [44]
g. This LR+ divides the retrained table's apparent sensitivity (6.2%, 30/482) by the cross-validated false-positive rate. This should be considered as an upper-range estimate.

By cross-validation, the mean sensitivity across folds was 4.4% with a 95% confidence interval of 1.7% to 7.1%. However, this is likely an underestimate and the in-sample sensitivity calculated directly was 6.2%. These performance estimates correspond to a positive likelihood ratio (LR+) of 18.5 (using sensitivity 4.4%) and 26.5 (using sensitivity 6.2%), meaning that a positive test result is about 26.5 times more likely in a mother with a child with autism than in a mother with a TD child. **(Table 3)**

### Distribution of predicted risks

The final test table is a binary predictor; but a continuous risk gradient can be derived by progressively scaling all thresholds in the table downward (by factors of 1.00, 0.99, 0.98, …, 0.01) and recording the resulting true positive and false positive calls at each scaling level. The resulting distribution of true and false positives is presented in **Figure 2**. Most notable in this visualization is the cluster of high-risk true positives, which is separated from the remainder of autism cases and all controls. Aside from this high-risk cluster which consisted largely of what has previously been defined as “transcendents” [29], the distribution of results across autism and typically developing mothers are not distinguishable (data not shown). The significance of this cluster of 30 true positives was assessed using the binomial test and it is highly significant (p value < 0.001). The analysis demonstrates that, as predicted risk increases, the proportion of autism and TD participants are similar for predicted risk below 100%. However, at 100% all predicted high-risk participants have autism. A small number of participants, including one mother of a TD child, had a predicted risk of approximately 95%. They first tested positive when the thresholds were lowered slightly, the point at which control mothers began to exceed them. This underscores the value of threshold inflation by providing an additional buffer against false positives.

**Figure 2.**
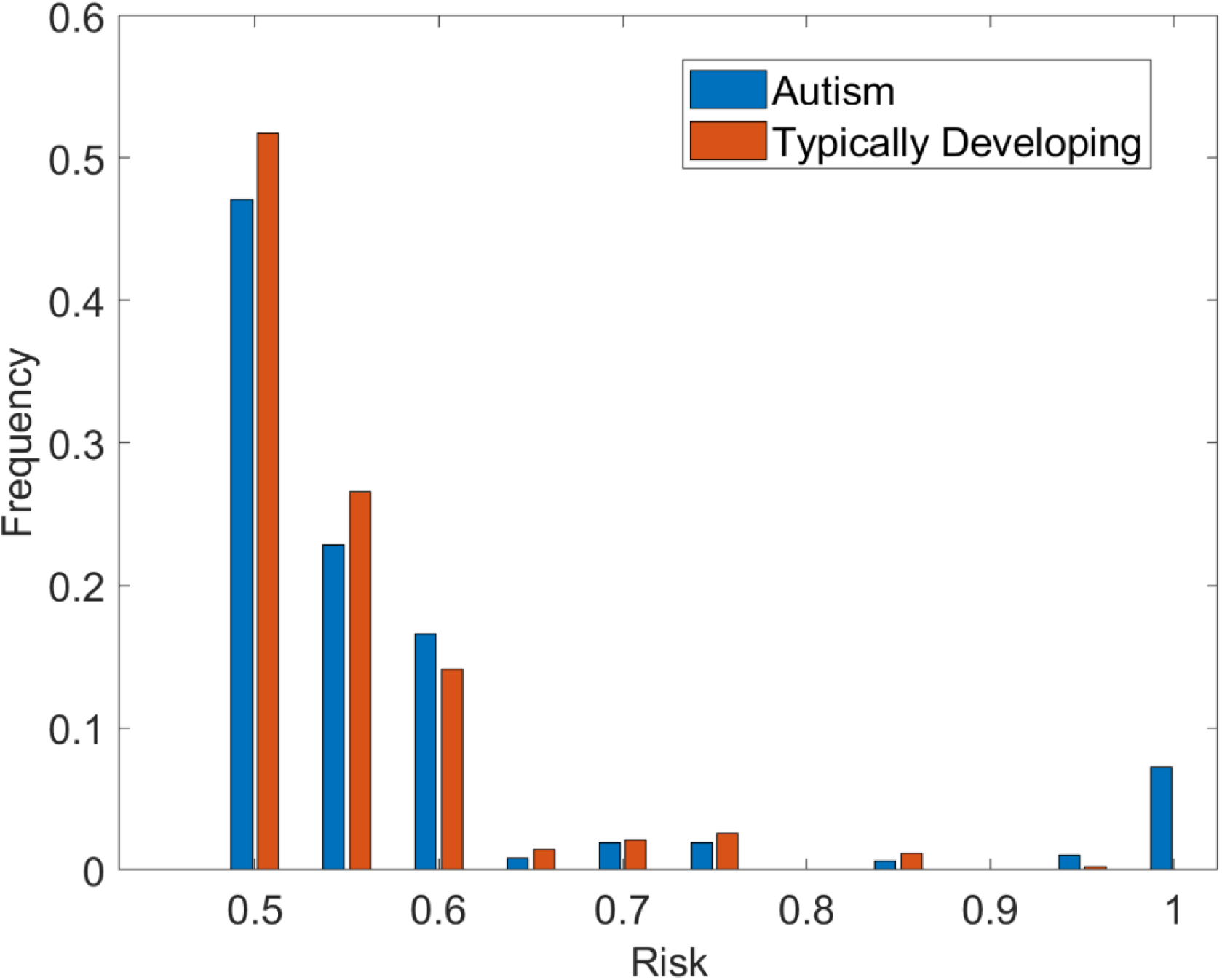
Distribution of predicted risk for mothers of autistic and TD children (combined CHARGE and MARBLES cohorts) Predicted risk was obtained by progressively scaling all thresholds of the test table generated by the cross-validation procedure downward (factors of 1.00 to 0.01 in steps of 0.01) and calculating the PPV at each level. Bars show the proportion of mothers of autistic children (blue) and TD children (orange) at each level of predicted risk, normalized within each group.

### Permutation Testing

Across 10,000 permutation iterations, 1.5% produced test tables containing five or more autoantibody combinations, compared to the five combinations selected in the final test table. Only 6 of 10,000 permutations produced 21 or more screen-positive cases, compared to the 21 screen-positive cases produced by the final test table in cross-validation (p = 0.0007).

### Performance metrics across methodologies

Overall, these methodologies provided robust performance estimates for the final test table from the 20-fold cross validation. Estimates from an external validation on the MARBLES cohort (42 mothers of children with autism and 90 mothers with TD children), reported in our sister publication, are also included [29]. **Table 3** summarizes the performance estimates across these datasets. Specificity and PPV remain consistent across methodologies with NPV varying only due to prevalence.

## Discussion

Since its first description nearly 20 years ago, MARA has gained considerable support as a biologically defined subset of autism, and in a series of studies culminating in this report, we have translated these findings into a clinical-grade test for autism risk [28,29]. In this study, we developed a clinical test table for the MARA autoantibody assay using the combined CHARGE and MARBLES cohorts (907 mothers) and estimated its performance by stratified 20-fold cross-validation. Five autoantibody combinations (CRMP-1 + CRMP-2, LDH-A + STI1, LDH-A + YB-1, LDH-B + YB-1, and STI1 + YB-1) were selected in 15 to 18 of the 20 folds and formed the final test table. By cross-validation, the test table achieved a specificity of 99.8% and an imputed mean PPV of 97.5% (95.5% pooled across folds), corresponding to a positive likelihood ratio of 18.5. The detection rate was 4.4% by cross-validation and 6.2% for the final test table in the full cohort. These results met the prespecified performance goals of this study. Permutation testing confirmed that the number of test-positive autism cases exceeded that expected by chance (p = 0.0007). Together, these findings support the MARA autoantibody test as a rule-in test that identifies a subset of high-risk mothers whose children have a very high probability of autism.

The selected combinations agree closely with our prior work, indicating that they reflect reproducible associations rather than chance features of a particular training sample. In the discovery and external validation of transcendent mothers [29], five combinations accounted for 60 of 70 transcendent cases in CHARGE and 9 of 14 in MARBLES. Four of these (CRMP-1 + CRMP-2, LDH-A + STI1, LDH-B + YB-1, and STI1 + YB-1) were selected here, with LDH-A + YB-1 replacing CRMP-2 + YB-1. Several of these combinations were also among the MAR autism-specific patterns previously reported with the prototype assay [13,20]. The emergence of the same combinations from prototype-assay studies, from discovery with the clinical assay, and from cross-validated test table development supports their role as robust markers of MARA, although these analyses drew largely on the same CHARGE population (see Limitations).

These performance estimates correspond to a likelihood ratio (LR+) of between 18.5 and 26.5 depending on whether using sensitivity directly from cross-validation or the derived best estimate from the final test table in sample analysis. **(Table 3)**This surpasses the standard interpretive framework of Deeks and Altman (2004), and indicates a substantial increase in the probability of autism following a positive test result [45]. As described by Deeks and Altman, an LR+ of 10 represents a large and often conclusive diagnostic shift, while an LR+ of approximately 30 represents a very large shift, moving a positive result from clinically meaningful to near-definitive rule-in across the range of clinically relevant pre-test probabilities. This incremental value in defining risk suggests this multiplexed ELISA assay appears to have potential for clinical implementation in a narrow clinical use case setting to identify a small subset of already high-risk mothers whose biomarker pattern implies very high autism risk, rather than maximizing sensitivity for all autism cases. That framing explains why the optimization was driven by near-perfect specificity and very high PPV, with detection rate across all autism only of secondary importance as this is a distinct subset of autism with a specific etiology.

Observed sensitivity in the small external validation (reported elsewhere) was higher (33.3%) based in part on the difference in the a priori risk basis of having an affected older sibling, and possibly the small size of that cohort [29]. Additionally, this highlights that any estimated value for sensitivity is potentially underestimated as there is no orthogonal method to test for MARA, i.e., clinically or with other biomarkers. Failure to recognize this limitation misses the significance of this as a distinct subset of autism.

Several design choices prioritized specificity over detection rate, consistent with the intended use. Cross-validation was used so that reported performance reflects participants not used to develop the test table; the final test table was then developed on the full cohort, but performance claims are based on cross-validation. Twenty folds were used so that each training partition contained 95% of participants, maximizing the number of controls available to set thresholds. Because the number of control mothers was modest, thresholds were also deliberately inflated to reduce the likelihood of false positives from control values higher than any observed in this cohort. These choices reduced the detection rate relative to our prior discovery work, in which 16.7% of mothers of autistic children in CHARGE were transcendent [29].

Transcendence was defined in-sample, with each case compared directly against all controls and without fixed thresholds, whereas the test table applies a small set of fixed, inflated thresholds to all participants and is limited to the five most contributory combinations. Mothers who were transcendent only on other combinations, or whose levels fell just below the inflated thresholds, are therefore not detected. The difference between the cross-validated (4.4%) and full-cohort (6.2%) detection (sensitivity) rates is likewise expected: threshold development on smaller training partitions and the small number of positives in each held-out fold both tend to reduce cross-validated detection. We consider this a reasonable trade-off for a rule-in test, in which a false-positive result carries considerable clinical and personal consequences, especially in preconceptual clinical practice. Potential refinement to the detection rate will be further addressed in a large forthcoming independent external validation study that is currently enrolling.

The clinical test for MARA discussed herein, based on the presence of one or more analytically validated autoantibody combinations targeting proteins critical to the developing brain, is intended to provide information as to whether a child or intended offspring is in fact at a very high likelihood of autism, compared to their already elevated risk based on their early clinical signs, or a mother having had a prior autistic child. Clinical application may enable earlier diagnosis and inform potential therapeutic or preventative strategies for this subset of autism.

Today it is generally believed that very early behavioral interventions may alter the cascade of emerging autism even in presymptomatic children [46–49]. Based on its high predictive value in the high-risk setting, this autoantibody test, when positive, could be considered a “companion diagnostic” for early behavioral intervention. In particular, this test may be useful for identifying those at extremely high risk for autism who might benefit from very early behavioral intervention, and ultimately, in identifying those individuals most likely to benefit from emerging biological or pharmacological modalities.

A compelling recent prospective study from Australia showed that infants who received preemptive intervention had lower odds of meeting diagnostic criteria for autism (7%) than those who received usual care (21%) at age three years, with the number needed to treat of seven participants [46]. Based on similar past studies [46–49], current American Academy of Pediatrics recommended screening for developmental disabilities at 18 and 24 months of age, or formal evaluation when autism is clinically suspected, can identify children who are at increased likelihood of ultimately receiving an autism diagnosis [50]. In that context a positive MARA autoantibody test may make a “watch-and-wait” approach undesirable for the family and the child.

Mothers who already have a child diagnosed with autism, who are desiring another child, face an empiric recurrence risk of approximately 20% [51]. They may also benefit from the test and its high predictive value of 97.5% when positive for pre-conceptual informed decision making [52]. While this may further support the decision to opt for reproductive stoppage after an affected child as nearly 30% actually do [53], families might elect surrogacy to avoid exposure to the offending autoantibodies, or implement potentially preemptive very early intervention in a subsequent child now expected to be at very high risk. It is vital to emphasize that the MARA autoantibody assays are not presently validated for the evaluation of an ongoing pregnancy, and the physiological and biochemical changes of pregnancy will need to be carefully evaluated.

Novel treatments to block the transfer of potentially harmful autoantibodies in the mother from reaching the fetus or even eliminate them in the mother altogether are under investigation. Congenital heart block in lupus [52], neonatal myasthenia gravis [52,53] and hemolytic disease of the fetus and newborn [54,55] are well known examples of diseases of infants arising through the same mechanism as proposed for MARA, and a class of monoclonal antibodies which block the FcRN receptor which mediates the transfer of maternal antibodies to the fetus [8,56] are under evaluation for such applications. A clinical trial for use in pregnancy for hemolytic disease of the fetus and newborn (HDFN) and fetal-neonatal alloimmune thrombocytopenia (FNAIT) is now in Phase III [54,57]. While IVIG has been broadly and safely used with varying benefits in relevant settings, including prenatally [55,56,58–64], preliminary results with FcRN blockers suggest they may be more effective, however further evaluation of both strategies will be needed. Additional means to lower or eliminate maternal autoantibodies capable of causing fetal disease include other classes of emerging agents which can selectively target and degrade these autoantibodies [8,9,56,65]. One study has demonstrated the ability to lower one of the MARA specific autoantibodies in an animal model [66]. In the future, testing to identify MARA could potentially become a companion diagnostic for such pharmaceutical interventions not just behavioral ones.

One limitation of the current study is that, although the samples were analyzed independently with an analytically validated clinical assay, the CHARGE population is the same as that used in our discovery study [29] and in previous academic studies conducted with the prototype “feasibility” assay [13,20]. Agreement across these analyses therefore supports the reproducibility of the findings with the clinical assay, but is not fully independent confirmation.

Developing the final test table on the full cohort could be misunderstood as inflating performance if its apparent full-cohort results were used as the principal evidence. This concern is addressed by separating test table development from performance evaluation: the final test table was developed using all participants, but performance claims are based on cross-validation, in which each participant was evaluated by a test table developed without that participant. Cross-validation nevertheless remains an internal estimate and does not replace validation in an independent population.

Combining two cohorts with somewhat different high-risk participants requires that they be sufficiently similar for a pooled analysis. Both CHARGE and MARBLES represent clinically higher-risk maternal intended-use populations, and the consistent selection of the same combinations across cross-validation folds of the pooled cohort supports their combination for this purpose. Performance may nonetheless differ between these settings, and the number of test-positive cases was too small to estimate performance separately in each.

The conservative design of the test table, including threshold inflation and restriction to the five most frequently selected combinations, means that some mothers at risk of having a child with MARA will not be detected. In the current clinical context, the resulting very high specificity, which limits the potential for false positives, is a reasonable trade-off. Because test-positive cases were few, estimates of detection rate and LR+ are also imprecise, as reflected in their wide confidence intervals. Further data generated from a current large external validation study will allow these estimates to be refined, and may allow a future version of the test to include additional combinations and capture a greater proportion of mothers whose children are affected by MARA autoantibodies.

PPV and NPV depend on the prevalence of autism in the population tested and will change when the test is applied to populations with different rates of autism. We report PPV and NPV at the study-level prevalence of the two high-risk cohorts in which the test was developed. These cohorts closely approximate the intended-use populations, but the 53% prevalence may not reflect all clinical settings. We therefore report here the study level PPV in the context of these high-risk clinical populations, rather than evaluating unsupported subgroup-specific recalculated PPVs, which should be prospectively recalibrated for each subsequent intended-use population once its prevalence estimates are firmly identified. These two cohorts do however closely approximate the intended-use groups for which the clinical assay is currently validated. The available data support the need for subgroup-specific recalibration in future work, but they do not provide sufficiently established subgroup prevalence estimates or an independent external validation dataset to calculate reliable, broadly applicable subgroup-specific PPV and NPV. Future studies will determine performance characteristics in populations with different autism prevalences to determine the test’s applicability across a broader range of clinical settings.

A related limitation is the absence of an independent reference standard for MARA. Because no other clinical or laboratory marker currently identifies this subset, sensitivity could be calculated only against all autism cases, and the test’s sensitivity for MARA itself cannot yet be determined. Similarly, the NPV is not low and a negative result should not be used to provide reassurance, consistent with the test’s intended use as a rule-in test.

In conclusion, using robust analytical approaches, the performance of the MARA autoantibody test was estimated by cross-validation, with results consistent with findings across previous studies. The performance characteristics of this assay support clinical implementation to identify a subset of mothers at very high risk for autism in their offspring, with potential as a companion diagnostic for interventions.

## Data Availability

All data produced in the present study are available upon reasonable request to the authors

## Author Contributions

Conceptualization, E.R.W.; Methodology, ERW, JvW, M.M.; Validation, M.M., J.S., Formal Analysis, M.M., E.R.W.; Investigation, ERW, M.M.; Resources, J.S., J.VdW., and ERW. Data Curation, M.M. and E.R.W.; Writing – Original Draft Preparation, E.R.W.; Writing – Review & Editing, M.M., J.VdW., E.R.W., M.P.; Project Administration, M.M., E.R.W., M.P., and J.VdW.; Funding Acquisition, M.M., E.R.W., M.P., and J.VdW.

## Institutional Review Board Statement

All materials used in this study used existing *de-identified* data/specimens where the investigator did not have access to private identifiable information and no contact with the subjects. Biospecimens used in this study were obtained in accordance with the Declaration of Helsinki, and approved by the Institutional Review Board of the University of California, Davis Protocol ID# 226028, Revised 02/26/2024, and all participants provided informed consent prior to inclusion in the study.

## Informed Consent Statement

Informed consent for participation was obtained from all subjects involved in the study at the originating institutions, but the use here was under waived approval for de-identified specimens. The study adheres to the scope of research allowed by the original consent.

## Data Availability Statement

The raw data supporting the conclusions of this article will be made available by the authors on request.

## Acknowledgments

The authors would like to thank: MARAbio’s investors including parents of children on the autism spectrum who sought to understand the biological cause of their child’s autism, participants in the UC Davis CHARGE and MARBLES studies, Drs. Mags McInerney and Jessica Barlow for assay development and laboratory services, Stuart Liu-Mayo for review of statistical methodology, and Dana Barberio of Edge Bioscience Communications for her contribution in editing the manuscript.

## Abbreviations

ELISA: Enzyme-linked immunosorbent assay
LDH-A: L-lactate dehydrogenase A chain
LDH-B: L-lactate dehydrogenase B chain
GAH: Guanine deaminase or cypin
STI1: Stress-induced phosphoprotein-1
NSE: Neuron-specific enolase
YB-1: Y-box-binding protein 1
CRMP-1: Collapsin response mediator protein 1
CRMP-2: Collapsin response mediator protein 2
LOB: Limit of Blank
LOD: Limit of Detection
LOQ: Lower Limit of Quantification
CI: confidence interval
ACD: acid–citrate–dextrose
RF: rheumatoid factor
SLE: systemic lupus erythematosus
YI: Youden index
OD: optical density or absorbance
FcRn: neonatal fragment crystallizable (Fc)-receptors
RoC: Receiver operator curve
LDT: Laboratory Developed Test
SPR: Sample Positive Rate
Sens: Sensitivity, Spec = Specificity
PPV: Positive Predictive Value
LR+: Likelihood Ratio (Positive)
Th: Threshold
IU: Intended Use

